# The clinical and prognostic significance of Protein Arginine Deiminase 2 and 4 (PADI2 & PADI4) in colorectal cancer

**DOI:** 10.1101/2020.08.14.20174920

**Authors:** Mohamed Gijon, Rachael L Metheringham, Michael S Toss, Samantha J Paston, Lindy G Durrant

## Abstract

**Aims:** Protein arginine deiminase (PADs) are a family of enzymes that catalyse the post translational modification (PTM) of proteins. Association between PAD expression with clinicopathology, protein expression and outcome was determined.

**Methods:** PADI2 and PADI4 expression was assessed immunohistochemically in a cohort of CRC patients.

**Results:** CRC tissues expressed variable levels of PADI2 which was mainly localised in the cytoplasm and correlated with patient survival (*p*=0.005); high expression increased survival time from 43.5 to 67.6 months. Expression of cytoplasmic PADI2 correlated with expression of nuclear β catenin, PADI4 and alpha-enolase. In contrast expression of nuclear PADI2 correlated with a decrease in survival (*p*=0.010), with high expression decreasing survival from 76.4 to 42.9 months. CRC tissues expressed variable levels of PADI4 in both the nucleus and cytoplasm. Expression of cytoplasmic PADI4 correlated with survival (*p*=0.001) with high expression increasing survival time from 48.1 to 71.8 months. Expression of cytoplasmic PADI4 correlated with expression of, nuclear β catenin, alpha-enolase (*p*≤0.0001, *p*=0.002) and the apoptotic related protein, Bcl-2. Expression of nuclear PADI4 also correlated with survival (*p*=0.011) with high expression of nuclear PADI4 increasing survival time from 55.4 to 74 months. Expression of nuclear PADI4 correlated with p53, alpha-enolase and Bcl-2. Multivariate analysis showed that TNM stage, cytoplasmic PADI2 and PADI4 remained independent prognostic factors in CRC. Both PADI2 and PADI4 are good prognostic factors in CRC.

**Conclusions:** High expression of cytoplasmic PADI2, PADI4 and nuclear PADI4 were associated with an increase in overall survival.

## INTRODUCTION

Colorectal cancer (CRC) is the third most common diagnosed cancer worldwide Despite substantial improvements in the diagnosis and treatment of CRC it still has a relatively low survival rate which estimated as 50-65% within 5 years depending on the healthcare systems [1]. The early detection of CRC could improve the outcome; however, early stage CRC often do not exhibit symptoms or have non-specific symptoms such which are usually misdiagnosed with other medical conditions [2]. However, most colon cancer patients will be diagnosed with regional or distant metastasis, thus adjuvant chemotherapy after surgery or palliative chemotherapy will often be needed [3].

The majority of CRCs (85%) arise from adenomas [4], the identification of the primary tumour, *BRAF* gene mutation status and tumour microsatellite instability (MSI) need to be determined to help guide clinician on the best treatment [5], which often being determined according to the progression of the disease, timing of the prior therapy, mutation profile and the toxicity profile of the corresponding drugs [6].

Approximately 4-5% CRC patients have tumours that are microsatellite instability-high (MSI-H) or mismatch repair deficient (dMMR), although these patients have a poor response to chemotherapy [7] they do have a clinical benefit following treatment with immune checkpoint inhibitors of PD-1/PD-L1 or in combination ipilimumab [8], [9]. A new approach for categorising CRC is to generate the Immune Score, which is defined by the number of lymphocytes infiltrating or surrounding the tumour. This score appears a strong prognostic factor for disease free and overall survival particularly in early-stage cancers. It also provides a tool or a target for novel therapeutic approaches, including immunotherapy [10]. T cell infiltration was a biomarker of good prognosis in CRC [11].

PAD enzymes catalyse the post translational modification (PTM) of proteins, in a process called citrullination. The exact biological function of citrullination remains obscure. Citrullination can also occur as a result of a degradation and recycling process called autophagy, and is induced in stressed cells [12], including cancer cells. Citrullination is catalysed exclusively by the family of Ca^2+^ dependent PAD enzymes that convert positively charged arginine residues within substrate proteins to neutrally charged citrulline, this modification can impact the protein structure, induce changes that result in protein denaturation and potentially alter the function of the modified proteins [13], [14]. Arginine deimination is recognised to be central for translational regulation of gene expression; to initiate tumour degradation, for formation of neutrophil extracellular traps (NET), and presentation of antigens within stressed cells to the immune system. Five highly conserved PAD enzymes, (1-4 and 6) have been reported in mammals, each exhibiting tissue specific distribution, subcellular localisation and substrate specificity.

PADI2 and PADI4 have been associated with several type of cancers such as CRC, breast or lung playing a critical role in cancer cells migration [15]–[19]. Both have widespread distribution and are the predominant types in immune cells [20]. The substrate for PADI2 includes the intermediate filament protein vimentin that is expressed by all mesenchymal cells, myelin basic protein (MBP), actin and keratins [21]. It has been shown that PADI2 negatively regulates β-catenin signalling [22]. β-catenin is critical for tumour development and is commonly activated in CRC. PADI4 can be translocated to the nucleus to citrullinate histones and some other transcription factors [23], [24]. PADI4 acts mainly as a transcriptional co-regulator; its substrates include multiple proteins involved in gene regulation of the *p53* pathway such as *ING4* [25], *p300* [26], *p21* [27] and *histones* (*H3, H2A and H4*) [28]–[30]. Other substrates for PADI4 includes nucleophosmin [31] and nuclear lamin C [32], both involved in apoptosis. Increased expression of PADI2 and PADI4 has been detected in malignant tumours, suggesting citrullination has induced tumourigenesis [33]. To date the prognostic value of PADI2 and PADI4 in CRC remains unclear. We have hypothesised that PADI2 and PADI4 are associated with a good outcome in CRC through their role in the citrullination process. Herein, we have investigated the prognostic value of PADI2 and PADI4 expression in a large cohort of CRC.

## PATIENTS AND METHODS

### Study cohort

This study was conducted on a cohort of CRC (n=247) from patients undergoing elective surgical resection of histologically proven primary CRC at Nottingham University Hospitals, Nottingham between January 1994 and December 2000. Written informed consent was obtained for all patients in accordance with the Declaration of Helsinki, ethical approval was obtained from the Nottingham Research Ethics committee (Q1020402). The AJCC cancer staging system was used (https://www.cancer.gov/about-cancer/diagnosis-staging/staging), where 3 TNM parameters are described and then TNM combinations are grouped into five less-detailed stages (stage 0-IV). For the analysis of clinicopathological variables for cytoplasmic and nuclear PADI2 and PADI4 expression the patient cohort was grouped into stages I-II and III-IV to allow the comparison of early tumours verses larger and metastatic tumours. Tumours histological subtypes were merged into two groups to obtain more informative statistical details. These groups comprised mucinous adenocarcinoma and adenocarcinoma which included all other carcinoma subtypes. The clinical details of the study cohort have been previously described [11], [36] and are summarised in Table 1. Clinicopathology information was obtained from pathology reports. Patients with lymph node positive disease routinely received adjuvant chemotherapy with 5-fluorouracil and folinic acid. The mean follow up period was 42 months (range 1-116). Disease specific survival (DSS) was determined from the date of resection of the primary tumour until December 31^st^ 2003 when any survivors were censored.

**Table 1:**
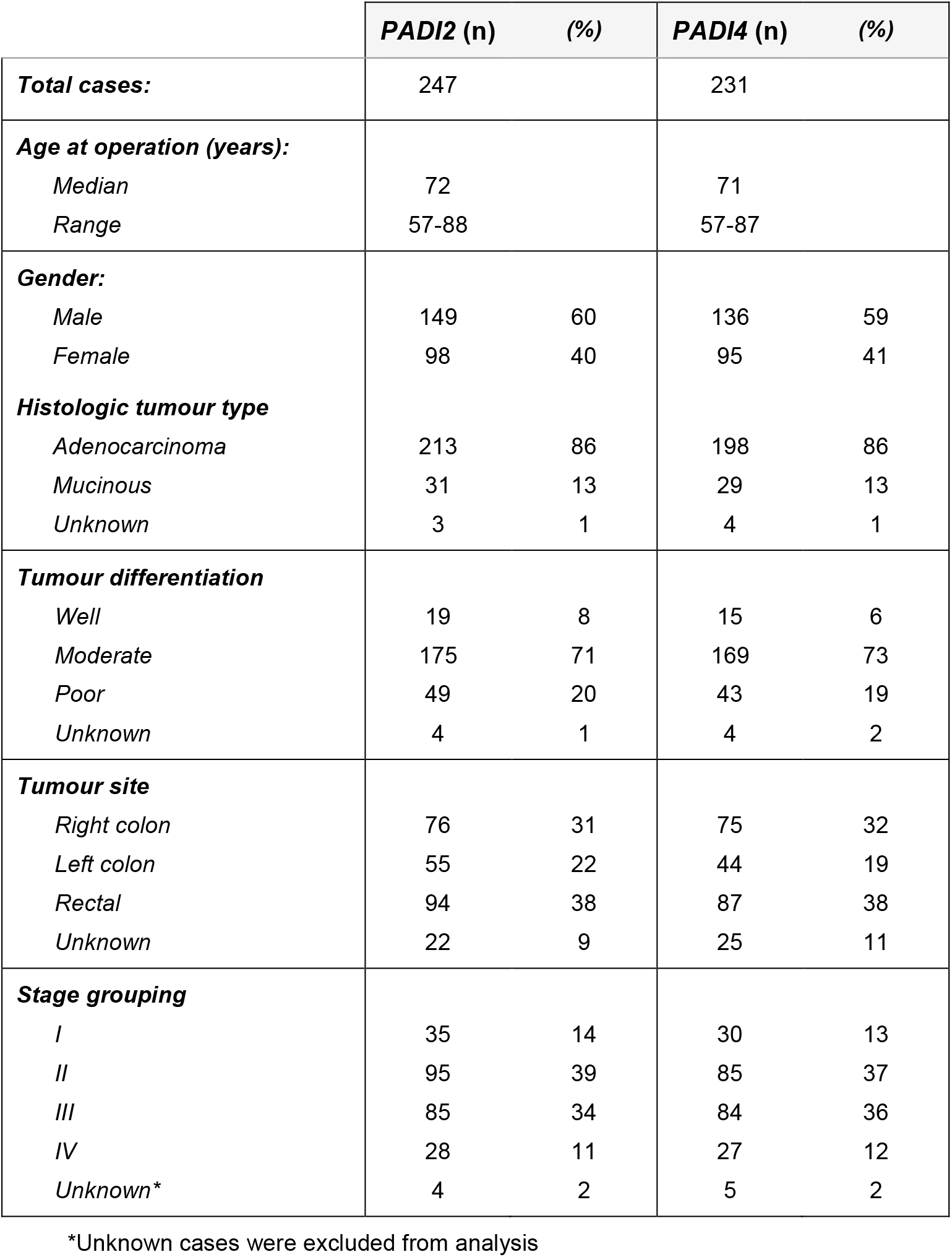
Clinicopathological details of the study cohort

### Immunohistochemistry (IHC)

Formalin fixed paraffin embedded (FFPE) tissue samples of the study cohort were retrieved from tissue archive where 4um thick sections were cut and stained with haematoxylin and eosin (H&E) stains for tumour burden assessment. Tissue microarray (TMA) was constructed from these samples prior to immunohistochemistry (IHC). To validate the TMAs for immuno-phenotyping, full-face sections of 40 randomly selected cases were stained and the protein expression levels were compared. The concordance between TMAs and full-face sections was compared using Cohen’s kappa test and reliability was excellent (kappa=0.8). IHC staining was performed using a streptavidin–biotin peroxidase method in conjunction with a Leica autostainer XL for tissue dewaxing and rehydration and the Novolink polymer detection system kit (Leica Biosystems, Newcastle-upon-Tyne, UK). Full face and TMA sections were first deparaffinised with xylene, rehydrated through graded alcohol and immersed in Peroxidase block (Novolink) for 5 minutes. The antigen retrieval was performed by immersing sections in citrate buffer at pH 6.0 and heating in a 800W microwave for 10 minutes at high power followed by 10 minutes at low power. Endogenous avidin/biotin binding was blocked (Protein blocking kit, Novolink), followed by addition of 100 μL Protein Block (Novolink) for 5 minutes. Sections were incubated for 1 hour with polyclonal rabbit anti-PADI2 antibody (Abcam, cat no 16478, 1:150 dilution), monoclonal rabbit anti-PADI4 antibody (Abcam cat no ab128086 1:700 dilution) for a 1 hour incubation at room temperature, monoclonal mouse anti-HLA-Class-II (DP, DQ, DR antigen) (Dako, cat no M0775, 1:1000 dilution) for 45 minutes at room temperature, monoclonal mouse anti-Vimentin antibody (Dako, cat no M7020, 1:150 dilution) for a 1 hour incubation at room temperature or monoclonal rabbit anti-ENO1 (Abcam, cat no ab155955, 1:100 dilution) for a 1 hour incubation at room temperature. The rest of the markers (Table 3) were stained in previous studies (P53 and Bcl2 [37], β-catenin [38], Ki-67, VEGFA and CEA [39]). After washing with tris buffered saline (TBS), sections were incubated with 100 μL of post primary antibody (Novolink kit) for 30 mins. After washing in TBS sections were incubated with 100 μL Novolink polymer for 30 mins. Sections were washed in TBS. Visualisation was achieved using 3, 3′-diaminobenzidine tetra hydrochloride (DAB, Novolink), and then counterstained with haematoxylin (Novolink), dehydrated in alcohol, cleared in xylene (Genta Medica, York, UK), and mounted with distyrene, plasticiser and xylene (DPX–BDH, Poole, UK).

**Table 2.** Clinicopathological variables for the patient cohort stained for cytoplasmic and nuclear PADI2 and PADI4

| **PADI2** | **Cytoplasmic PADI2 (%)** |  |  | **Nuclear PADI2 (%)** |  |  |
| --- | --- | --- | --- | --- | --- | --- |
| **Variable** | **Low PADI2** | **High PADI2** | **χ<sup>2</sup> test (p Value)** | **Low PADI4** | **High PADI4** | **χ<sup>2</sup> test (p Value)** |
| **Age**<br>≤ 72<br>> 72 | 22 (16.2)<br>20 (18.0) | 114 (83.8)<br>91 (82.0) | 0.702 | 18 (43.9)<br>15 (46.9) | 23 (56.1)<br>17 (53.1) | 0.800 |
| **Gender**<br>Male<br>Female | 24 (16.1)<br>18 (18.4) | 125 (83.9)<br>80 (81.6) | 0.644 | 20 (51.3)<br>13 (38.2) | 19 (48.7)<br>21 (61.8) | 0.264 |
| **Histologic tumour type**<br>Adenocarcinoma<br>Mucinous | 34 (16.0)<br>6 (19.4) | 179 (84.0)<br>25 (80.6) | 0.634 | 20 (50.0)<br>13 (20.0) | 19 (50.0)<br>21 (80.0) | 0.264 |
| **Tumour differentiation**<br>Well<br>Moderate<br>Poor | 2 (10.5)<br>28 (16.0)<br>10 (20.4) | 17 (89.5)<br>147 (84.0)<br>39 (79.6) | 0.586 | 3 (42.9)<br>24 (50.0)<br>6 (35.3) | 4 (57.1)<br>24 (50.0)<br>11 (64.7) | 0.571 |
| **Tumour site**<br>Right colon<br>Left colon<br>Rectal | 14 (18.4)<br>10 (18.2)<br>15 (16.0) | 62 (81.6)<br>45 (81.8)<br>79 (84.0) | 0.898 | 10 (45.5)<br>9 (60.0)<br>12 (42.9) | 12 (54.5)<br>10 (40.0)<br>16 (57.1) | 0.544 |
| **Stage grouping**<br>I - II<br>III - IV | 20 (15.4)<br>22 (18.8) | 110 (84.6)<br>95 (81.2) | 0.475 | 24 (57.1)<br>9 (29.0) | 18 (42.9)<br>22 (71.0) | **0.017\*** |
| **PADI4** | **Cytoplasmic PADI4 (%)** |  |  | **Nuclear PADI4 (%)** |  |  |
| **Variable** | **Low PADI2** | **High PADI2** | **χ<sup>2</sup> test (p Value)** | **Low PADI4** | **High PADI4** | **χ<sup>2</sup> test (p Value)** |
| **Age**<br>≤ 72<br>> 72 | 35 (27.1)<br>29 (28.4) | 94 (72.9)<br>73 (71.6) | 0.827 | 60 (46.5)<br>51 (50.0) | 69 (53.5)<br>51 (50.0) | 0.598 |
| **Gender**<br>Male<br>Female | 34 (25.0)<br>30 (31.6) | 102 (75.0)<br>65 (68.4) | 0.272 | 62 (25.0)<br>49 (31.6) | 74 (54.4)<br>46 (48.4) | 0.370 |
| **Histologic tumour type**<br>Adenocarcinoma<br>Mucinous | 56 (28.3)<br>6 (20.7) | 142 (71.7)<br>23 (79.3) | 0.391 | 93 (47.0)<br>16 (55.2) | 105 (53.0)<br>13 (44.8) | 0.370 |
| **Tumour differentiation**<br>Well<br>Moderate<br>Poor | 4 (26.7)<br>44 (26.0)<br>14 (32.6) | 11 (73.3)<br>125 (74.0)<br>29 (67.4) | 0.691 | 7 (46.7)<br>77 (45.6)<br>25 (58.1) | 8 (53.3)<br>92 (54.4)<br>18 (41.9) | 0.336 |
| **Tumour site**<br>Right colon<br>Left colon<br>Rectal | 24 (32.0)<br>11 (25.0)<br>21 (24.1) | 51 (68.0)<br>33 (75.0)<br>66 (75.9) | 0.498 | 41 (54.7)<br>21 (47.7)<br>36 (41.4) | 34 (45.3)<br>23 (52.3)<br>51 (58.6) | 0.240 |
| **Stage grouping**<br>I - II<br>III - IV | 29 (25.2)<br>35 (30.2) | 86 (74.8)<br>81 (69.8) | 0.400 | 50 (43.5)<br>61 (52.6) | 65 (56.5)<br>55 (47.4) | 0.166 |

**Table 3.** Univariate analysis of cytoplasmic and nuclear PADI2 and PADI4 expression in correlation with tumour markers or treatment using the χ2 or Fisher’s exact test.

| Variable | PADI2 cytoplasmic | PADI2 nuclear | PADI4 cytoplasmic | PADI4 nuclear |
| --- | --- | --- | --- | --- |
| | $\chi^2$ test<br>(p Value) | $\chi^2$ test<br>(p Value) | $\chi^2$ test<br>(p Value) | $\chi^2$ test<br>(p Value) |
| PADI2 cytoplasmic |  | 0.394 | **<0.0001\*\*** | **0.048\*** |
| PADI2 nuclear | 0.394 |  | 0.145 | 0.336 |
| PADI4 cytoplasmic | **<0.0001\*\*** | 0.145 |  | **<0.0001\*\*** |
| PADI4 Nuclear | **0.048\*** | 0.336 | **<0.0001\*\*** |  |
| MHC class II in tumour | 0.407 | 0.476 | 0.398 | 0.370 |
| Vimentin | 1.000 | 0.094 | 0.529 | 0.554 |
| α-Enolase cytoplasmic | **0.001\*\*** | 0.729 | **<0.0001\*\*** | 0.469 |
| α-Enolase nuclear | 0.088 | 0.670 | **0.002\*\*** | **0.011\*** |
| Distant metastasis | 0.687 | 0.658 | 0.059 | 0.143 |
| Nuclear β-catenin | **0.020\*** | 0.988 | **0.004\*\*** | 0.549 |
| P53 | 0.645 | **0.024\*** | 0.922 | **0.016\*** |
| Ki-67 | 0.368 | 0.474 | 0.224 | 0.765 |
| Bcl2 | 0.975 | 0.764 | **0.032\*** | **0.022\*** |
| VEGFA | **0.001\*** | 0.933 | 0.363 | 0.267 |
| carcinoembryonic antigen (CEA) | **0.042** | 0.243 | 0.522 | 0.091 |
| Chemotherapy | 0.840 | 0.500 | 0.846 | 0.599 |
| Radiotherapy | 0.938 | 0.351 | 0.424 | 0.123 |

\*\* Correlation is significant at the 0.01 level (2-tailed).
\* Correlation is significant at the 0.05 level (2-tailed).

### Slide scanning, viewing and scoring

TMA stained slides were digitalised using NanoZoomer (Hamamatsu, Bridgewater, NJ, USA) high throughput scanner. Images were visualised using Aperio ImageScope v11.2.0.780 software. Cytoplasmic PADI2 and cytoplasmic and nuclear PADI4 proteins expression was assessed using the percentage of positivity (0-100%) and staining intensity (Negative: Score 0, Weak: Score 1, Moderate: Score 2, and Strong: Score 3) following the Histo-score system (H-score 0 to 300. Negative=0; weak=1-100; moderate=101-200 and strong=201-300). Observers were blind to clinical and pathological parameters.

### Statistical analysis

Statistical analysis was performed using SPSS21 statistical software (SPSS Inc., Chicago, IL, USA). Pearson’s χ2-tests and Fisher’s exact tests were used to determine the significance of associations between categorical variables. The cohort was categorised into two groups by defining the optimal cut-off points for cytoplasmic PADI2 (H-score 150), nuclear PADI2 (H-score 160), cytoplasmic PADI4 (H-score 100) and nuclear PADI4 (H-score 260) expression by X-tile software (X-Tile Bioinformatics Software, Yale University, version 3.6.1). Survival rates were calculated using the Kaplan–Meier method; differences between survival curves were examined using the log-rank test. The Cox regression model was used for multivariate analysis in order to calculate the Hazard ratios and independent significance of individual factors. In all cases two-sided P-values of <0.05 were considered as statistically significant.

## RESULTS

### Frequency and distribution of PADI2 and PADI4 expression in the study cohort

After exclusion of uninformative cores (i.e., loss or folding of cores during tissue sectioning and processing), there was 247 and 231 cores suitable for scoring of PADI2 and PADI4 expression respectively. Clinicopathological details of each marker are described in Table 1.

In this cohort 99% of the cases expressed PADI2 in the cytoplasm, where 28% showed nuclear and cytoplasmic co-expression. PADI4 showed cytoplasmic and nuclear in all cases. Occasional surrounding inflammatory cells and fibroblasts showed weak expression to PADI2 and PADI4, however, this was not evaluated further in this study.

According to the H-score assessment. After categorisation of each marker using the optimal cut offs there was 62% of the tumours showed high expression of cytoplasmic PADI2, 28% moderate expression, 9% weak expression and only 1% were negative for PADI2, representative staining is shown in Figure 1A-C. For nuclear PADI2 30% tumours showed strong expression, 32% moderate expression, 34% weak expression and only 4% were negative for nuclear PADI2. The H-score assessment for cytoplasmic PADI4 showed all tumours expressed PADI4, 11% strong expression, 61% moderate expression and 28% weak expression, representative staining is shown in Figure 1D-F. All tumours expressed nuclear PADI4, 77% strong expression, 19% moderate expression and 4% showed weak expression.

**Figure 1.**
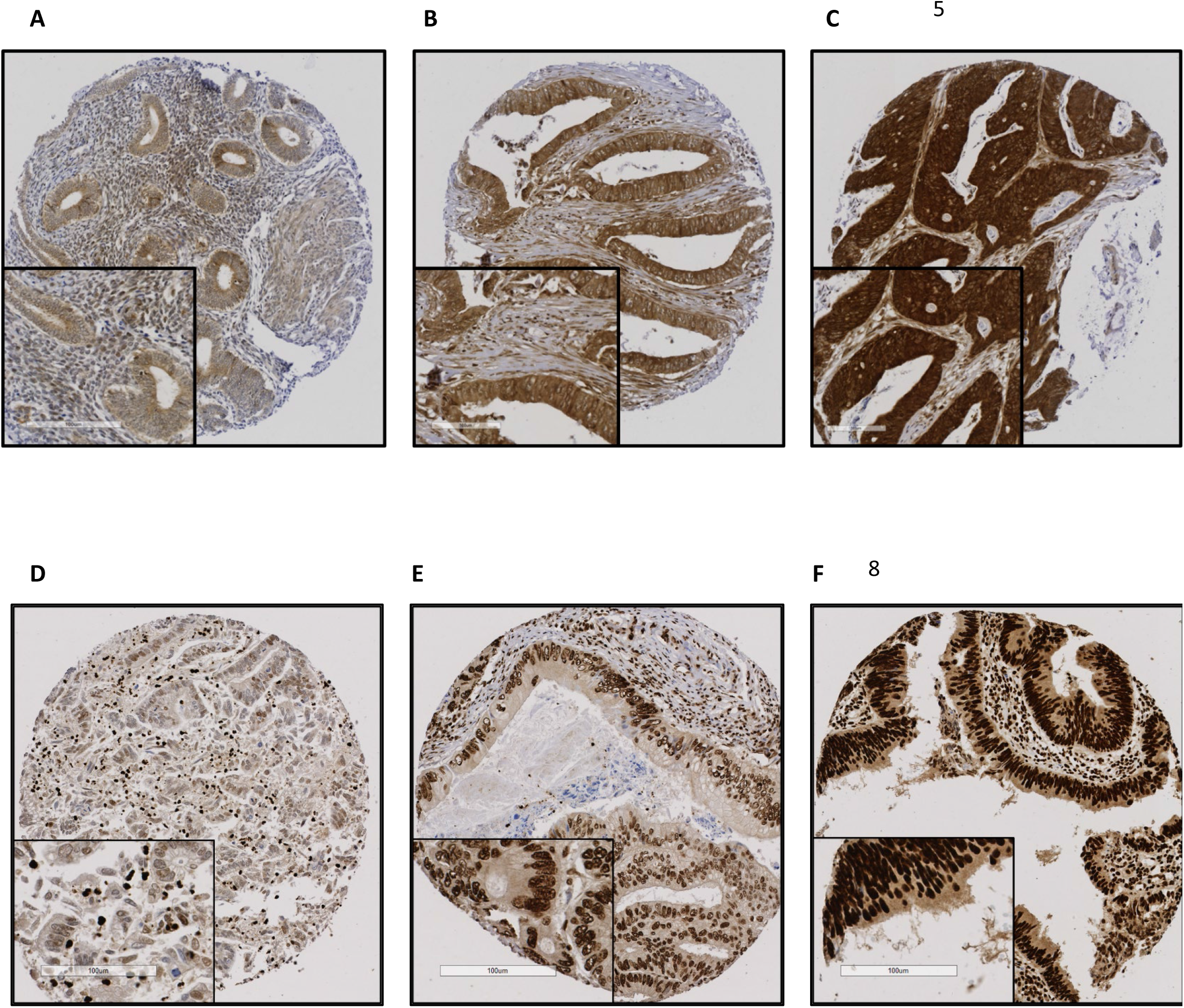
Representative immunohistochemistry staining of PADI2 (A-C) and PADI4 (D-F) in colorectal tissue microarray images. The level of expression ranged from weak (A, D), moderate (B, E) and strong expression (C, F) according to the H-score.

### Correlation of PADI2 and PADI4 expression with the clinicopathological variables

High nuclear expression of PADI2 was significantly correlated with the TNM stage (*p*=0.017), while no significant association were observed between nuclear PADI4, cytoplasmic PADI2 or PADI4 expression with any of the other clinicopathological parameters (Table 2).

The results from the univariate analysis for cytoplasmic PADI2 with protein expression showed that there is a significant correlation with nuclear β-catenin (*p*=0.020), vascular endothelial growth factor A (VEGFA) (*p*=0.001) and carcinoembryonic antigen (CEA) (*p*=0.042), cytoplasmic and nuclear PADI4 (*p*≤ 0.0001, *p*=0.048) and cytoplasmic alpha-enolase (*p*≤ 0.001). The results from the analysis for nuclear PADI2 showed only a correlation with the tumour suppressor protein, p53 (*p*=0.024). The results from the univariate analysis for cytoplasmic PADI4 showed there is a significant correlation with nuclear β-catenin (*p*=0.004), nuclear PADI4 (*p*≤ 0.0001), cytoplasmic and nuclear alpha-enolase (*p*≤ 0.0001, *p*≤0.002) and the apoptotic related protein, Bcl-2 (*p*=0.032). The results from the analysis for nuclear PADI4 showed there is a significant correlation with nuclear alpha-enolase (*p*=0.011), p53 (*p*=0.016) and Bcl-2 (*p*=0.022) (Table 3).

### PADI2 and PADI4 expression and outcome

High cytoplasmic expression of PADI2 showed a significant correlation with better survival (*p*=0.005) (Figure 2A). In contrast, high nuclear expression of PADI2, (Figure 2B) was associated with a decrease in survival (*p*=0.010). Higher cytoplasmic and nuclear PADI4 (Figures 2C and 2D) showed there was a significant correlation with survival with tumours expressing cytoplasmic PADI4 and nuclear PADI4 (*p*=0.001 and *p*=0.011, respectively).

**Figure 2.**
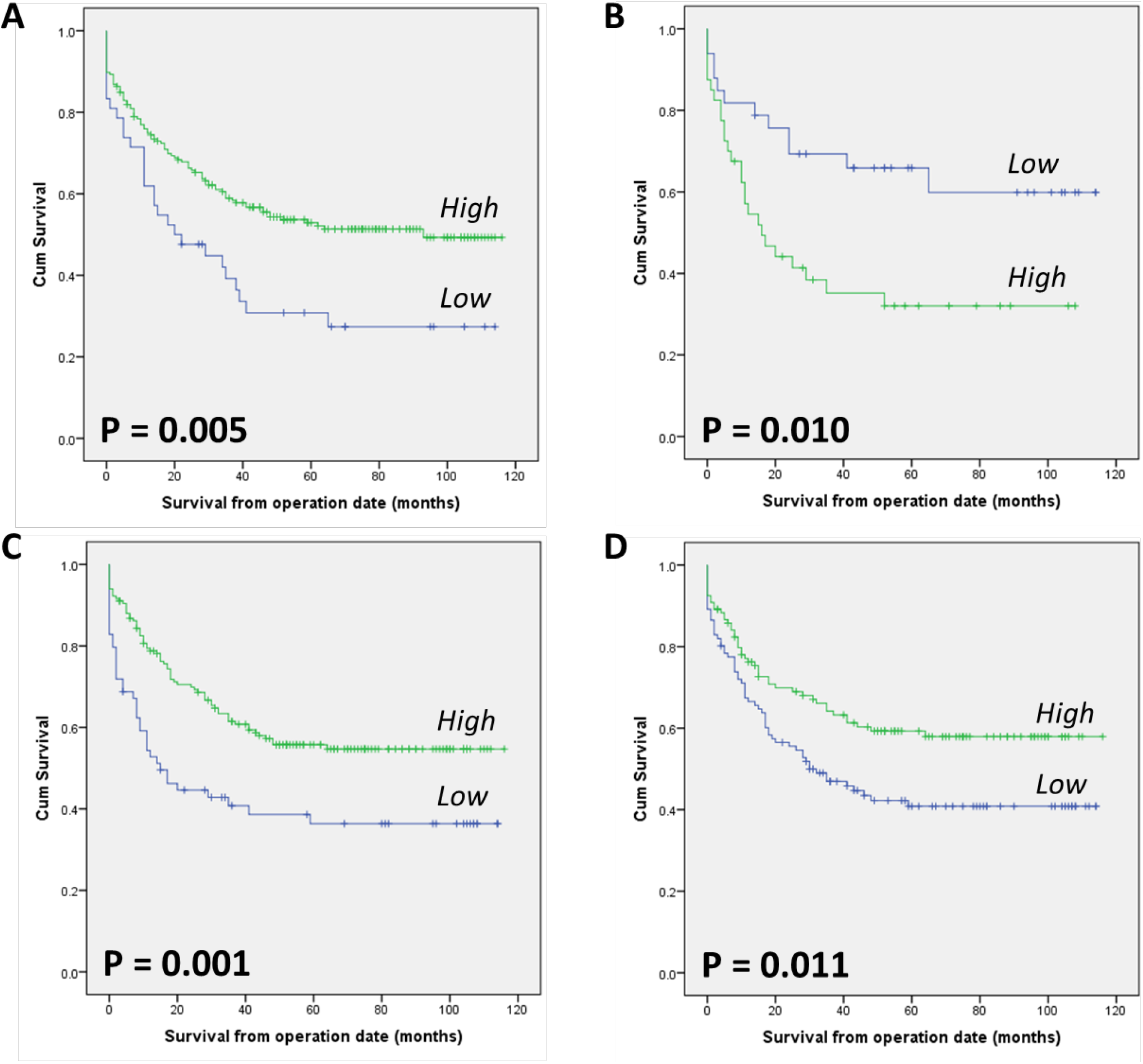
Kaplan-Meier curves for disease survival for CRC patients from the time of tumour resection showing high (green line) or low expression (blue line) of PADI2 or PADI4, (A) cytoplasmic PADI2, low = 42 cases, high = 205 cases (B) nuclear PADI2, low = 33 cases, high = 40 cases (C) cytoplasmic PADI4, low = 64 cases, high = 167 cases (D) nuclear PADI4, low = 111 cases, high = 120. Significance determined using log-rank test.

### Cytoplasmic PADI2 and PADI4 expression by colorectal tumours are independent prognostic markers

Multivariate Cox regression model including other factors known to be associated with poor outcome in CRC (TNM stage and vascular invasion), showed high cytoplasmic expression of PADI2 (Table 4) was an independent prognostic protective marker (*p*<0.001). The expression of cytoplasmic PADI4 (Table 4) also conferred independent tumour protection.

**Table 4.** Multivariate analysis results (Cox regression model) for TNM stage, Vascular Invasion, Nuclear β-catenin and cytoplasmic and nuclear PADI2 and PADI4 expression.

| PADI2 |  | Cytoplasmic PADI2 |  |  | Nuclear PADI2 |  |  |
| --- | --- | --- | --- | --- | --- | --- | --- |
| VARIABLE | CATEGORY | HAZARD RATIO (HR) | 95% CI | P VALUE | HAZARD RATIO (HR) | 95% CI | P VALUE |
| Stage | I and II<br>III and IV | 2.290 | 1.689 to<br>3.105 | **<0.001\*** | 2.651 | 1.590 to<br>4.421 | **<0.001\*** |
| Vascular Invasion | Negative<br>Positive | 1.437 | 0.937 to<br>2.204 | 0.097 | 2.152 | 0.934 to<br>4.957 | 0.072 |
| PADI2 (Cytoplasmic) | Low<br>High | 0.458 | 0.288 to<br>0.728 | **0.001\*** | N/A |  |  |
| PADI2 (nuclear) | Low<br>High | N/A |  |  | 1.677 | 0.755 to<br>3.722 | 0.204 |
| PADI4 |  | Cytoplasmic PADI4 |  |  | Nuclear PADI4 |  |  |
| VARIABLE | CATEGORY | HAZARD RATIO (HR) | 95% CI | P VALUE | HAZARD RATIO (HR) | 95% CI | P VALUE |
| Stage | I and II<br>III and IV | 2.238 | 1.613 to<br>3.104 | **<0.001\*** | 2.303 | 1.658 to<br>3.200 | **<0.001\*** |
| Vascular Invasion | Negative<br>Positive | 1.493 | 0.953 to<br>2.339 | 0.080 | 1.435 | 0.917 to<br>2.247 | 0.114 |
| PADI4 (Cytoplasmic) | Low<br>High | 0.629 | 0.398 to<br>0.994 | **0.047\*** | N/A |  |  |
| PADI4 (nuclear) | Low<br>High | N/A |  |  | 0.727 | 0.469 to<br>1.126 | 0.153 |

Significant p values are in bold

## DISCUSSION

To improve the survival in patients with CRC its early diagnosis and treatment is critical. A number of prognostic makers have been investigated to identify patients at high risk of relapse. The identification of a reliable prognostic marker has been challenging. Post translational modifications of the human proteome can have a major impact on health and disease, increasing the functional diversity of proteins by adding new functional groups. There is accumulating evidence that citrullination is implicated in cancer development and can be an excellent target to alert the immune response to recognise and remove stressed cells including cancer cells. This study aimed to investigate the prognostic value of PADI2 and PADI4 in CRC.

PAD enzymes are activated by a high concentration of calcium, typically in the millimolar range [12]. In healthy viable cells the PAD enzymes are maintained in an inactive state due to the low concentrations of Ca^2+^ [13], in double membrane vesicles within viable cells the calcium concentration can be high leading to the activation of the PAD enzymes. The PAD enzymes play a key role in the regulation of gene expression through the citrullination of histones and transcription factors and have been shown to have a role in tumour progression [21]. Citrullination can occur within autophagosomes as a result of autophagy, here the activated PAD enzymes citrullinate engulfed proteins from the cytoplasm [40], [41], this process is induced in stressed cells [42] including cancer cells. During inflammation, epitopes are citrullinated and can be subsequently presented by MHC class II (MHC-II) molecules for recognition by CD4 T cells [42]. A number of studies in autoimmune patients have demonstrated that CD4 T cell responses can be detected to citrullinated proteins such as vimentin and enolase [43]–[46]. We have already shown that citrullinated α-enolase and vimentin are excellent targets for anti-tumour immunity inducing potent immune responses [43], [47], [48] and *in vivo* tumour rejection [49].

Our data shows that 99% of CRC tumours express PADI2 and all tumours in our cohort expressed PADI4. Increasing evidence suggests that the PAD enzymes play a role normal cell transformation and tumour progression. We also show that in the majority of cases PADI4 was expressed in both the cytoplasm and nucleus, whereas PADI2 was mainly located in the cytoplasm with only 28% of tumours showing nuclear expression. Analysis of the clinicopathological parameters for the cohort stained with PADI2 and PADI4 revealed that only nuclear PADI2 showed a correlation with a clinical parameter, notably TNM stage. Both PADI2 and PADI4 target different cellular substrates [50], [51], defined by the cell type and the subcellular localisation of the substrate, this indicates that PADI2 and PADI4 acts as an independent prognosis markers.

We investigated if the expression of PADI2 or PADI4 correlated with other markers involved in cancer, cell survival and progression. Nuclear PADI2 correlated with p53 expression and both conferred a worse prognosis [37]. In contrast the expression of nuclear PADI4 conferred a good prognosis and inversely correlated with p53 expression. Anwar *et al*. analysed the results from 35 studies where p53 was found to predict of a worse outcome [52]. Multiple interactions between PADI4 and p53 have been reported, suggesting the importance of PAD-induced citrullination during apoptosis. The expression of the major p53 target gene OKL38 was suppressed by the p53-mediated recruitment of PADI4 to the promoter of OKL38 and the subsequent removal of histone arginine methylation mark, thus directly modulating apoptosis [53]. PADI4 was found to bind and then citrullinate the inhibitor of growth 4 (ING4), another tumour suppressor protein that is known to bind to p53. The PADI4 mediated citrullination of ING4 at the nuclear localisation sequence region prevents p53-to-ING4 binding, suppresses p53 acetylation, and subsequently inhibited downstream p21 expression [25]. Nuclear and cytoplasmic PADI4 conferred a good prognosis and correlated with expression of Bcl-2 which is also a predictor of good prognosis [37]. Despite the defined role of Bcl-2 protein in suppression of apoptosis, in CRC the evidence suggests that Bcl-2 expression is correlated with favourable parameters [54] and a better prognosis [55]. Although Bcl-2 inhibits apoptosis it has also been shown to suppress tumour cell growth [56] and better differentiation of the tumours, possibly resulting in a more favourable outcome. It has been reported that Bcl-2 contains an anti-proliferative domain, distinct from the domains required for its anti-apoptotic activity [57], and also Bcl-2 can be converted to a BAX-like death effector by cleavage of a regulatory loop domain by caspases [58]. Cytoplasmic PADI2 and PADI4 expression correlated with nuclear β-catenin expression and both correlated with good prognosis in this cohort. Wnt/β-catenin signalling is important for carcinogenesis of certain tumours, and PADI2-mediated citrullination was recently linked to this pathway [22]. The anti-parasitic drug temozolomide (NTZ) inhibited Wnt/β-catenin signalling through directly targeting PADI2 as demonstrated by co-immunoprecipitation studies. The addition of NTZ resulted in a significant increase of β-catenin citrullination by PADI2 and also substantially extended the half-life of PADI2 protein, suggesting that NTZ enhances the stability of PADI2. Furthermore, the results from colony growth assays have suggested that PADI2-mediated citrullination of β-catenin could limit the proliferation of CRC cells by inhibiting the Wnt pathway. As a result, this elegant study uncovered a hitherto unrecognised mechanism of Wnt signalling inhibition via PADI2.

The PAD enzymes have been described as playing a role in early tumour cell transformation and tumour progression. In addition, the PAD enzymes also catalyse the PTM of proteins under conditions of cellular stress, in the process of citrullination. Citrullination occurs as a result of the degradation and recycling process of autophagy. In the presence of inflammation, epitopes are citrullinated and can be subsequently presented by MHC-II molecules for recognition by CD4 T cells [42]. We have recently shown that citrullinated α-enolase and vimentin are excellent targets for anti-tumour immunity inducing potent immune responses [43], [47], [48] and *in vivo* tumour rejection [49]. It was therefore of interest that cytoplasmic PADI2 correlated with cytoplasmic and nuclear alpha-enolase, and cytoplasmic PADI4 correlated with vimentin and cytoplasmic and nuclear alpha-enolase suggesting that these may be good targets for citrullination in colorectal cancer, these tumour cells can then be recognised and removed by the immune system.

In conclusion, our results show that high expression of cytoplasmic PADI2, PADI4 and nuclear PADI4 were associated with an increase in overall survival. Nuclear expression of PADI4 may confer a good prognosis by regulating expression of genes such as the p53/Bcl-2. Cytoplasmic PADI2 and or PADI4 may confer a good prognosis by altering cell signalling pathways such as via β-catenin or may be involved in protein degradation which can lead to presentation of citrullinated epitopes for detection by cytotoxic CD4 T cells. Cancer vaccines targeting citrullinated vimentin and/or enolase maybe a good therapy for CRC patients expressing high levels of PAD enzymes [12].

## Data Availability

All data relevant to the study are included in the article or uploaded as supplementary information. Data is available upon request.

## Acknowledgements

The authors would also like to thank Dr Tina Parsons for critical reading of the manuscript, Maria Diez and Chris Nolan for their technical support to this research.

## Statement of authors contributions

MG developed the methodology, performed experiments, interpreted IHC staining data and collected clinical data. LD, MG, RM performed statistical analysis. LD contributed to the study design. MG wrote the draft manuscript. MT, SP, LD provided critical review of the data and contributed to the final manuscript. LD supervised the study. All the authors reviewed and approved the final manuscript.

